# High Seroprevalence of anti-SARS-CoV-2 antibodies among Ethiopian healthcare workers

**DOI:** 10.1101/2021.07.01.21259687

**Authors:** Tesfaye Gelanew, Berhanu Seyoum, Andargachew Mulu, Adane Mihret, Markos Abebe, Liya Wassie, Baye Gelaw, Abebe Sorsa, Yared Merid, Yilkal Muchie, Zelalem Teklemariam, Bezalem Tesfaye, Mahlet Osman, Gutema Jebessa, Abay Atinafu, Tsegaye Hailu, Antenehe Habte, Dagaga Kenea, Anteneh Gadissa, Desalegn Admasu, Emmet Tesfaye, Timothy A. Bates, Jote Bulcha, Rea Tschopp, Dareskedar Tsehay, Kim Mullholand, Rawleigh Howe, Abebe Genetu, Fikadu G. Tafesse, Alemseged Abdissa

**Affiliations:** Armauer Hansen Research Institute, Addis Ababa Ethiopia; Department of Medical Microbiology, School of Biomedical and Laboratory Sciences, College of Medicine and Health Sciences, University of Gondar, Gondar, Ethiopia; Arsi University, Asella College of Health Sciences, Asella, Ethiopia; Hawassa University, College of Medicine and Health Sciences, Department of Medical Microbiology, Hawassa, Ethiopia; All Africa Leprosy and Tuberculosis Rehabilitation and Training Center (ALERT Center) Hospital, Addis Ababa, Ethiopia; College of Health and Medical Sciences, Department of Medical Laboratory Sciences, Haramaya University, Harar, Ethiopia; Department of Molecular Microbiology & Immunology, Oregon Health & Sciences University, OR, USA; Horae Gene Therapy Center, University of Massachusetts Medical School, MA, USA; Swiss tropical and Public Health Institute, Basel, Switzerland; London School of Hygiene and Tropical Medicine, London, UK

**Keywords:** SARS-CoV-2, COVID-19, RBD, ELISA, seroprevalence, antibodies, Ethiopia

## Abstract

**Background:** COVID-19 pandemic has a devastating impact on the economies and health care system of sub-Saharan Africa. Healthcare workers (HWs), the main actors of the health system, are at higher-risk because of their occupation. Serology-based estimates of SARS-CoV-2 infection among HWs represent a measure of HWs’ exposure to the virus and a guide to the prevalence of SARS-CoV-2 in the community. This information is currently lacking in Ethiopia and other African countries. This study aimed to develop an in-house antibody testing assay, assess the prevalence of SARS-CoV-2 antibodies among Ethiopian high-risk frontline HWs.

**Methods and findings:** A cross-sectional seroprevalence study was conducted among HWs in five public hospitals located in different geographic regions of Ethiopia. Socio-demographic and clinical data were collected using questionnaire-based interviews. From consenting HWs, blood samples were collected between December 2020 and February 2021, the period between the two peaks of COVID-19 in Ethiopia. The collected sera were tested using an in-house immunoglobin G (IgG) enzyme-linked immunosorbent assay (ELISA) for SARS-CoV-2 specific antibodies on sera collected from HWs. Of 1,997 HWs who provided a blood sample, demographic and clinical data, 50.5% were female, 74.0% had no symptoms compatible with COVID-19, and 29.0% had history of contact with suspected or confirmed patient with SARS-CoV-2 infection. The overall seroprevalence was 39.6%. The lowest (24.5%) and the highest (48.0%) seroprevalence rates were found in Hiwot Fana Specialized Hospital in Harar and ALERT Hospital in Addis Ababa, respectively. Of the 821 seropositive HWs, 224(27.3%) had history of symptoms consistent with COVID-19. A history of close contact with suspected/confirmed COVID-19 cases was strongly associated with seropositivity (Adjusted odds Ratio (AOR) =1.4, 95% CI 1.1-1.8; p=0.015).

**Conclusion:** High SARS-CoV-2 seroprevalence levels were observed in the five Ethiopian hospitals. These findings highlight the significant burden of asymptomatic infection in Ethiopia, and may reflect the scale of transmission in the general population.

**Author summary:** *Why was this study done?:*

- Severe acute respiratory syndrome coronavirus 2 (SARS-CoV-2) is a global public health threat, including Africa
- The actual morbidity and mortality associated with SARS-CoV-2 infection in Ethiopia underestimated due to the limited molecular testing capacity.
- We have limited knowledge about the seroprevalence of COVID-19 among health workers in Ethiopia.
- This study aimed to develop an in-house immunoglobin G (IgG) enzyme-linked immunosorbent assay (ELISA) for SARS-CoV-2 specific antibodies on sera collected from HWs and to find out the proportion of healthcare workers who have developed antibodies specific to SARS-CoV-2 from five public hospitals located in the different regions of Ethiopia.

*What did the researchers do and find?:*

- A cross-sectional seroprevalence study was conducted among HWs in five public hospitals located in different geographic regions of Ethiopia.
- Socio-demographic and clinical data were collected from recruited and consented participants using questionnaire-based interviews.
- Blood samples were collected from participants between December 2020 and February 2021, the period between the two peaks of COVID-19 in Ethiopia.
- The collected sera were tested using an in-house ELISA for SARS-CoV-2 specific antibodies on sera collected from HWs.
- Approximately 40% of the 1,997 healthcare workers who participated in this study had antibodies against SARS-CoV-2 infection.
- No association between seropositivity and study participants’ age, gender, occupation, and comorbid medical conditions.

*What do these findings mean?:*

- The observed high seroprevalence among healthcare workers regardless of their occupation suggests the cryptic but massive SARS-CoV-2 transmission in urban hospital settings.
- Most of the seropositive healthcare workers in the present study were asymptomatic, and might pose a threat to the most vulnerable populations such as individuals with comorbid medical conditions.
- Given the low level of vaccine roll-out (1%), this study highlights the need to strengthen health workers’ adherence to personal protection practices such as wearing face masks to protect individuals at high risk of developing severe COVID-19 illness after SARS-CoV-2 infection.

## 1. Introduction

Despite the total population of 1.3 billion, Africa stands out as the region least affected by the Severe Acute Respiratory Syndrome-Corona-Virus-2 (SARS-CoV-2) and coronavirus disease-2019 (COVID-19) pandemic. As of May 23^rd^, 2021^1^, the total reported case number had risen to 4,748,581 with 128,213 reported deaths, representing 2.9% and 3.7% of global cases and deaths, respectively. The low number of reported cases and deaths in Africa have been attributed to low testing capacity, younger population, warmer environments, and the successful implementation of control measures^2^. Also, pre-existing cross-protective immunity due to the four other less pathogenic human coronaviruses (HCoVs)^3^, Bacillus Calmette-Guérin (BCG)-vaccination^4^, or recent history of malaria infection may offer some protection against infection or severe forms of COVID-19^5^.

To date, Ethiopia has performed over 2,682,758 real-time reverse transcription-polymerase chain reactions (RT-PCR) tests for SARS-CoV-2 and reported 268,901 cases and 4,068 deaths since the first case was detected in the country on March 13, 2020. Almost all testing have been done to confirm SARS-CoV-2 infection in suspected cases and contacts, as well as both outbound and inbound travelers. Given the difficulty and cost of RT-PCR-based testing in resource-limited countries like Ethiopia, mildly affected or asymptomatic individuals are not usually screened, and so the number of confirmed SARS-CoV-2 infections is likely vastly underestimated^6^. In this context, seroprevalence surveys are of the utmost importance to assess the proportion of the population that have already developed antibodies against the virus.

Evidence has shown that healthcare workers (HWs) are at higher risk of acquiring the infection than the general population. This is because their work is likely to require close contact with SARS-CoV-2 infected patients at COVID-19 treatment centers, in emergency rooms and wards, and via virus-contaminated surfaces. If infected, they can pose a significant risk to vulnerable patients and co-workers^8^. Thus, assessing the seroprevalence of SARS-CoV-2 antibodies among HWs in Ethiopia will help us understand COVID-19 spread among health care facilities and to measure the success of public health interventions. It will also provide an opportunity to compare the disease trajectory in a low-income setting. A report from London, UK suggested that the rate of asymptomatic SARS-CoV-2 infection among HWs reflects general community transmission rather than in-hospital exposure^9^. Therefore, a serosurvey of SARS-CoV-2 was conducted amongst HWs in five public hospitals to estimate the Seroprevalence of SARS-CoV-2 in urban Ethiopia. We then discuss the implications of our SARS-CoV-2 serosurveillance for frontline healthcare workers and the Ethiopian population at large.

## 2. Methods

### Participant recruitment

This cross-sectional study represents a joint effort between the Armauer Hansen Research Institute (AHRI) and five public hospitals in Ethiopia, namely Gondar, Asella, Hawassa, Hiwot Fana (located in Harar), and All Africa Leprosy and Tuberculosis Rehabilitation and Training Center (ALERT Center) hospitals. These participating hospitals were selected because they are among the 11 hospitals located in different regional states of the country, and are linked to the AHRI’s Clinical Research Network. Similar seerosurvey studies for the remaining hospitals liked to the AHRI’s CRN are ongoing. Ethical approvals were obtained from all institutions and written informed consent was obtained from each participant. All hospital staff (n=7,898) from all five public hospitals were invited to take part in the study through office memos and notice board announcements. However, only 24.4% of them were volunteered to provide 5 milliliter blood and demographic and clinical data. Demographic and clinical data were obtained using a structured questionnaire based on WHO SARS-CoV-2 seroprevalence studies (World Health Organization (WHO, the “Solidarity II” global serologic study for COVID-19)^10^.

### Sample collection, storage, transportation, and inactivation

Five milliliters of blood were collected in a serum collection tube from each participant using standard procedures. Sera were separated by centrifugation and stored at -20 °C until transferred to AHRI laboratory in Addis Ababa, Ethiopia, in a cold box. Inactivation of infectious viruses in serum was performed by incubation with Triton X-100 to a final concentration of 1% for 1 hour ^11^ and stored at - 80°C until testing for the presence of SARS-CoV-2 specific antibodies. Serum samples were collected from December 2020 to February 2021 between the two peaks of SARS-CoV-2 transmissions in Ethiopia (https://covid19.who.int/region/afro/country/et).

### Enzyme-Linked Immunosorbent Assay (ELISA)

The SARS-CoV-2 spike protein Receptor Binding Domain (RBD)-containing plasmid construct was cloned as described previously^12^. The RBD protein was then expressed in EXPi293 cells using previous methods^12^. Then, the purified RBD protein was used as a target antigen to develop our in-house anti-SARS-CoV-2 RBD IgG detection ELISA. We used 1µg/ml of RBD to coat the microwell plate overnight at 4°C. The assay is an indirect ELISA, measuring serum IgG against RBD of spike protein SARS-CoV-2, using a horseradish peroxidase-linked anti-human IgG secondary antibody (Invitrogen, USA).

Supplementary method shows the detail procedure description of our assay (Supplementary Method). We validated this ELISA using pre-COVID-19 pandemic sera/plasma samples (n=365), WHO “Solidarity II” plasma panels (n=5), and sera/plasma samples (n=401) collected from a cohort of mild (majority) and severe COVID-19 patients confirmed by RT-PCR. Detection of RBD-specific IgG antibodies in each serum sample was done in duplicate microwells of ELISA plate. In each ELISA run, we included positive and negative controls. Positive and negative control samples were selected by matching their optical density (OD) readouts with WHO solidarity II plasma panels developed by the United Kingdom’s National Institute for Biological Standards and Control (NIBSC;20/130, single donor, high-titer antibody), 20/120 (single donor, relatively high-titer antibody), 20/122 (pool of five donor samples, mid-titer antibody), 20/124 (low S1, high-nucleocapsid protein antibody titer), 20/126 (low-titer antibody, 20/128, negative control).

### Optimization and validation of in-house anti-RBD IgG detection ELISA

We noted background signal from the negative controls at a 1:100 dilution of serum. Of 365 pre-COVID-19 sera, 30 showed optical density (OD) values comparable to the low reactive convalescent WHO plasma samples. We further optimized the assay by increasing the concentration of skimmed milk powder and Tween-20 in blocking buffer from 3% to 4% and from 0.05% to 0.1%, respectively, and serum dilution at 1:200. Except for fourteen pre-COVID-19 samples, the background was significantly reduced when re-tested false positives, which in turn increased the specificity of our assay without compromising its sensitivity in WHO positive control samples and serum samples obtained from a cohort of COVID-19 patients.

Using our optimized ELISA protocol, we calculated the cut-off value for positivity using pre-COVID-19 pandemic sera collected between 2012 and 2018, and plasma/serum samples collected from cohort of confirmed COVID-19 patients at different time points of post-onset of symptoms (dps). The definition of seropositivity represents a greater than 2.5 ratio of sample OD value to the mean OD value of the negative controls (Fig 1). This definition provides specificity of 97.7% (95% CI, 95.6-99.0) (Table S1). Our anti-RBD IgG detection ELISA showed a sensitivity of 67.3% (95% CI 62.3.0-72.3), 75. 8% (95% CI 61.0-86.0), 100% (95% CI 84.0-100) in serum/plasma samples collected at 1-7 dps, 8-14 dps and ≥15dps, respectively from mostly (>90%) mild and moderate COVID-19 cases confirmed by RT-PCR (Table S2). This performance is in line with those published for both in-house and commercial assays approved for emergency use by the FDA^13^. and https://covid-19-diagnostics.jrc.ec.europa.eu/.].

**Figure 1.**
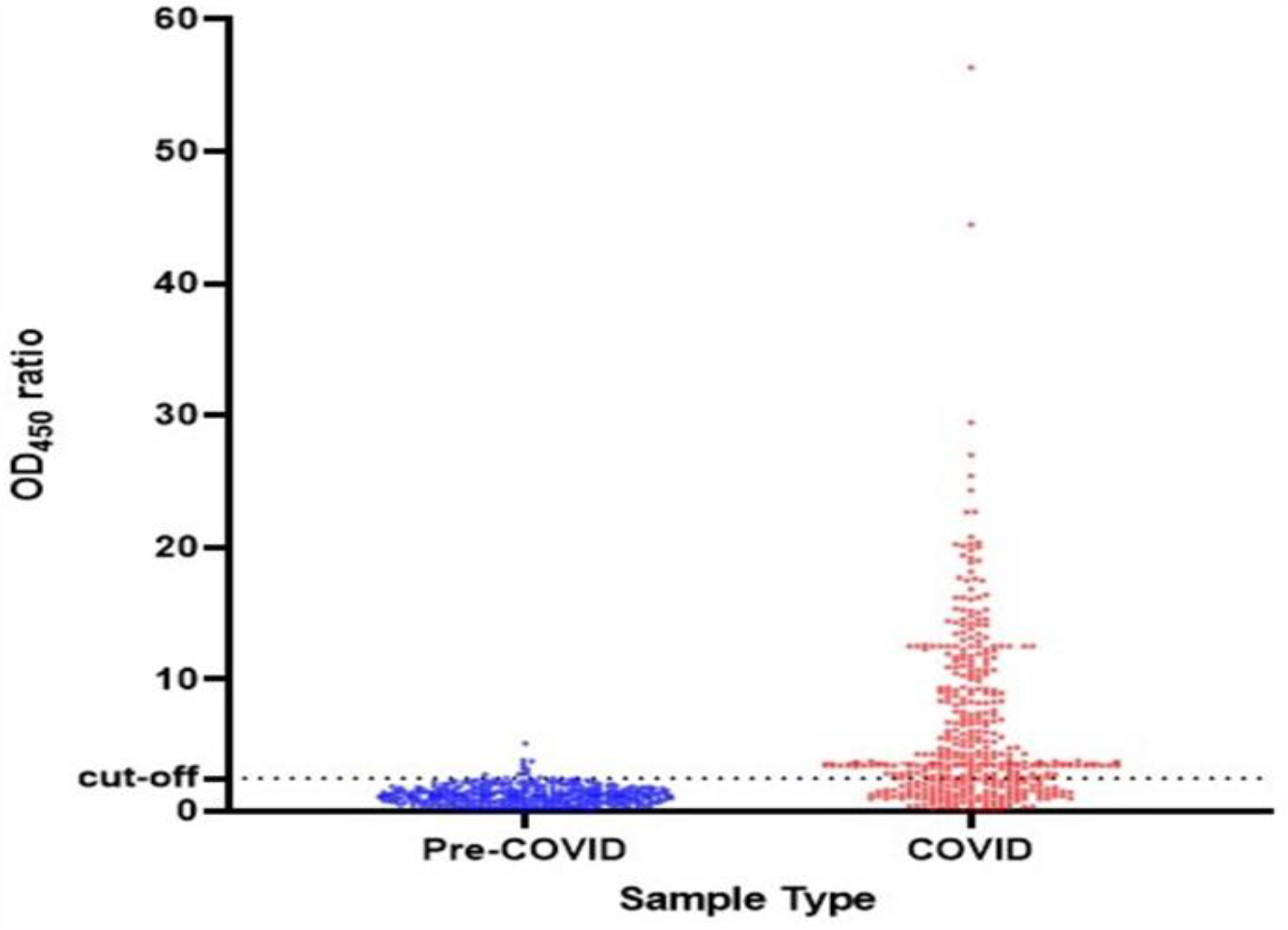
Validation of the SARS-CoV-2 RBD specific IgG antibody detection ELISA. The value on the y-axis represents the ratio of OD_450_ nm to the average mean OD_450_ nm of the negative controls. The broken black line represents the cut-off value (2.5). We tested a total of 405 serum/plasms samples collected from cohort of mild and moderate (93.6%) and severe Ethiopian COVID-19 patients confirmed by RT-PCR (represented in red color). Of these 325 samples were collected during 0-7 days post-onset of symptoms (dps); 52 were collected during 8-14 dps, and 17 were collected within 15-28 dps (Table S2). We also tested serum/plasma samples collected from 365 Ethiopian individuals before the global COVID-19 pandemic, represented in blue color (Table S1).

### In-house IgG ELISA comparison with commercial anti-SARS-CoV-2 serologic assays

We further compared the relative sensitivity and specificity of our assay with commercially available SARS-CoV-2 antibody tests: one lateral flow assay (LFA) (Hangzhou Realy Tech Co., LTD) and one ELISA (Beijing Wantai Biological Pharmacy Enterprise Co., Ltd) following the manufacturers’ instructions using randomly selected small panels (pre-pandemic; n=40, and COVID-19; n=40) from the large size panels that were used for our assay validation. We found a comparable sensitivity and specificity to those commercially available COVID-19 antibody detection kits depending on the sample collection date (Table S3 and Table S4). We then utilized this assay to estimate the seroprevalence of anti-SARS-CoV-2 spike protein RBD IgG antibodies among HWS.

### Data analysis

The data were double entered into REDCap Database Version 8.11. Following data verification and validation, analysis was done using STATA Version 15.0. Descriptive statistics and the actual number of cases were used to describe frequency outputs for categorical variables. Figures were generated using GraphPad Prism Version 9.1. Cross-tabulations were performed to explore and display relationships between two categorical variables. The overall seroprevalence with 95% CI for anti-SARS-CoV-2 RBD IgG was calculated by dividing the number of seropositive cases divided by the total number of study participants from all five hospitals. Apparent SARS-CoV-2 prevalence was stratified by the geographic location of hospitals, age, sex·, self-reported previous history exposure, symptoms, comorbidities, and further by occupation/department where HWs are working. Bivariate logistic regression was done between seroprevalence with independent variables such as sex, age, occupation, comorbidity, history of close contact, and symptoms. Multivariate regression analysis was applied for those variables with a p-value <0.25 in bivariate analysis to evaluate the strength of association between independent variables and seropositivity, the outcome variable. A p-value of <0.05 was considered statistically significant.

## 3. Results

### Characteristics of study participants

The total number of HWs in the five participating hospitals was 7,898. Of these, we enrolled 1,997 (24.4%) HWs [from Gondar (n=453); Assela (n=484); ALERT (n=308); Hawassa (n=414); and Haromaya (n=338)] in the study. Almost half (50.7 %) of the study participants were female. The majority (85.7%) of the participants belonged to the age groups 25-34 and 35-49 years with the mean age 34 years (range 20-60 years). Of the participants, 559 (28.3%) were nurses, 368 (18.5%) were doctors, 223 (11.3%) were medical laboratory personnel, 345 (17.5%) were administrative staff, and the remaining 24.2% (n=478) did not specify their occupation. In the cohort, 1490 (74.0%) participants were asymptomatic, 507 (26.0%) had reported one or more symptoms compatible with COVID-19 during the preceding 4 weeks, and 557 (29.0%) had a history of close contact with suspected or confirmed COVID cases. Overall, 133 (6.7%) of the participants reported having a history of comorbid medical conditions, with obesity (1.9%), asthma (1.7%), hypertension (1.5%), and Human Immunodeficiency Virus (HIV) (1.3%) being the most common. These and other demographic and clinical characteristics of study participants are summarized in Table 1.

**Table 1.** Participant characteristics with interview data (n=1,997) and seroprevalence, Ethiopia, 2021.

| Characteristics | N | % | Crude seroprevalence (%), 95% CI |
| --- | --- | --- | --- |
| **Gender** |  |  |  |
| Male | 980 | 49.3 | 39.6 (36.6-42.7) |
| Female |  | 51.7 | 42.37(39.4-45.5) |
| **Age (in years)** |  |  |  |
| 19-24 | 169 | 8.8 | 44.0(36.7-51.7) |
| 25-34 | 918 | 46.0 | 41.6(38.4-44.8) |
| 35-49 | 792 | 39.7 | 39.7(36.4-43.0) |
| ≥50 | 115 | 5.8 | 41.7(33.00-51.0) |
| **Morbidity** |  |  |  |
| Yes | 133 | 6.7 | 44.4(36.1-52.9) |
| No | 1864 | 93.3 | 40.9(38.7-43.3) |
| **COVID-19** |  |  |  |
| Symptomatic | 507 | 26.0 | 39.9(37.4-42.4) |
| Asymptomatic | 1490 | 74.0 | 45.2(40.9-49.5) |
| **Contact** |  |  |  |
| Yes | 557 | 29.0 | 48.5(44.3-52.6) |
| No | 1362 | 71.0 | 38.1(35.6-40.7) |
| **Hospitals** |  |  |  |
| ALRET | 308 | 15.4 | 48.1(40.3-53.6) |
| Asella | 484 | 24.2 | 40.7(36.4-45.1) |
| Gondar | 453 | 22.6 | 44.7(40.12-49.3) |
| Hawassa | 414 | 20.7 | 44.8(40.05-49.) |
| Hiwot Fana | 338 | 17.0 | 24.6(20.3-29.4) |
| **Occupation** |  |  |  |
| Doctor | 368 | 18.7 | 40.5(35.6-45.6) |
| Nurse | 559 | 28.3 | 41.9(37.7-45.8) |
| Lab Tech | 223 | 11.3 | 46.2(39.7-52.8) |
| Administrator | 345 | 17.4 | 39.1(34.1-44.4) |
| Others | 478 | 24.2 | 43.5(38.7-48.4) |

N is the total number of participants included in each category.
% indicates proportion of participants that fell within each category.

### Seroprevalence of SARS-CoV-2 antibodies by geographic locations of participating hospitals, age, sex, healthcare cadre and clinical factors

The overall seroprevalence of SARS-CoV-2 antibodies among HWs from all five studied public hospitals was 833 of 1,997 (39.6.7% [95% CI 40. 37.4-41.7]). The estimated seroprevalence with 95% CI for each of the participating hospitals was shown in Fig 2 and Table 1, ranging from 24.5% to 48.0%. We did not find association between seropositivity and participants’ demographic and clinical features given in Table 1, except history of contact with confirmed or confirmed COVID-19 contact. Non-significant seroprevalence difference was observed between females (42.4% [95% CI 39.4-45.55]) and males (39.6% [95% CI 36.6-42.7]). However, higher [48.5% [95% CI 44.3-52.6)] seroprevalence found in HWs who had close contact with COVID-19 case than in HWs who reported no contact (38.1% [95% CI 35.6-40.7]). Seroprevalence was similar amongst different cadres of the health system, and amongst different age groups of HWs (Table 1). Slightly higher (44.4% [95% CI 36.1-52.9]) seropositivity against SARS-CoV-2 was found in comorbid HWs than in HWs who had no comorbidity (40.9% [95% CI 38.7-43.3]).

**Figure 2.**
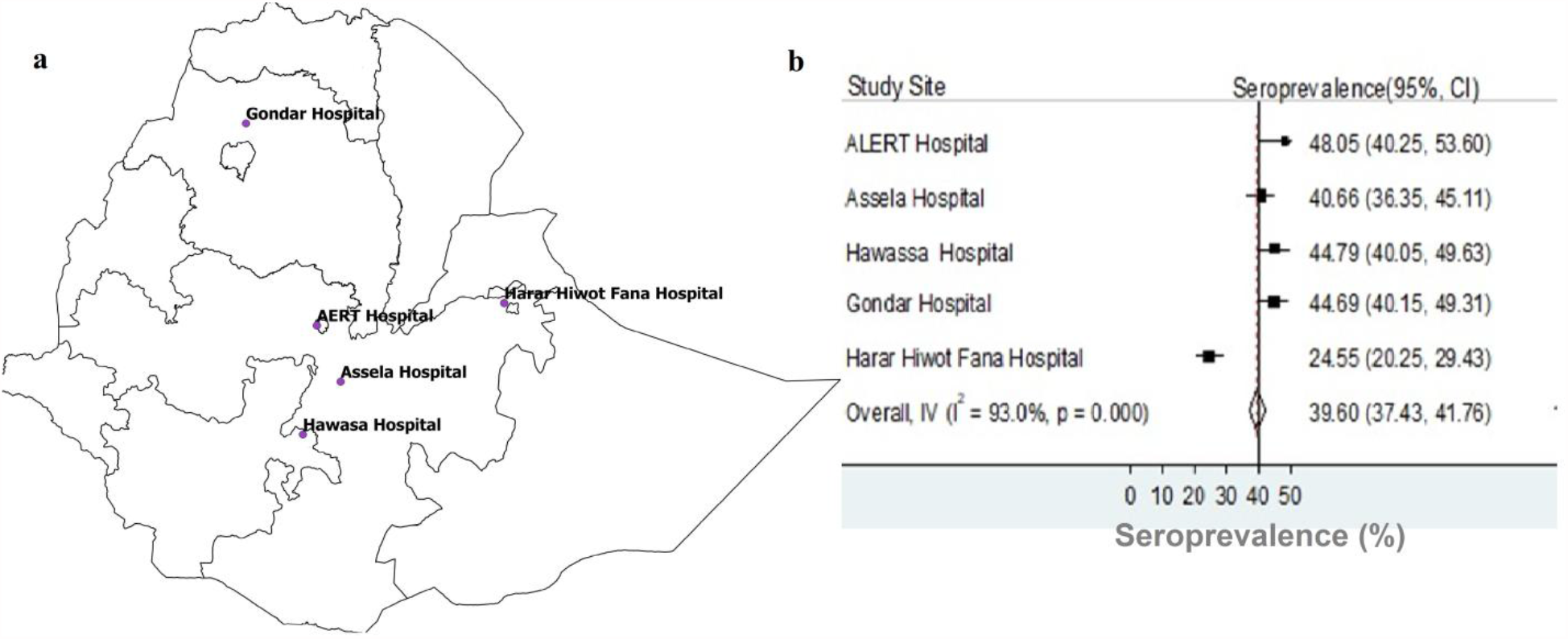
A map of Ethiopia showing the location of the five hospitals from which a total of 1997 healthcare workers enrolled between December 2020 and February 2021 (**a**), and the corresponding seroprevalence of severe acute respiratory syndrome coronavirus 2 (SARS-CoV-2) (**b**). The y-axis of Fig 2b represents the study hospitals. The x-axis of Fig 2b shows crude seroprevalence rates (%) with 95% confidence intervals estimated by dividing the number of participants tested seropositive for immunoglobin G (IgG) antibodies elicited against the receptor binding domain (RBD) of the spike protein of SARS-CoV-2 to the total number of participants who provided sera and were tested.

### Factors associated with anti-SARS-CoV-2 RBD IgG antibodies positivity

HWs working at Gondar (AOR=2.8, 95% CI 1.99-3.87; p=0.001), ALERT (AOR=2.7, 95% CI 1.6-3.1; p=0.001), Hawassa (Adjusted OR=2.1, 95% CI 11.5-3.2; p=0.001) and Assela (AOR=2.1, 95% CI 1.6-3.1; p=0.001) were at higher odds of seropositivity compared to HWs working at Hiwot Fana Specialized University Hospital (Table 2).

**Table 2.** Odds ratios (OR) of seropositivity by general characteristics of study participants, Ethiopia, 2021.

| Variable | Adjusted OR (95% CI) | p-value |
| --- | --- | --- |
| **Hospital** |  |  |
| Hiwot Fana | 1 |  |
| ALERT | 2.7(1.6-3.1) | **0.0001** |
| Assela | 2.2 (1.6-3.1) | **0.0001** |
| Gondar | 2.8(2.0- 3.9) | **0.0001** |
| Hawassa | 2.2 (1.5-3.2) | **0.0001** |
| **Sex** |  |  |
| Male | 1 |  |
| Female | 1.1(0.92-1.4) | 0.222 |
| **Age (in years)** |  |  |
| 19-24 | 1.27(0.9-1.9) | 0.226 |
| 25-34 | 1.1(0.9-1.5) | 0.254 |
| 35-49 | 1 |  |
| >=50 | 1.2(0.8-1.8) | 0.479 |
| **Contact** |  |  |
| No | 1 |  |
| Yes | 1.4 (1.1-1.8) | **0.015** |
| **COVID-19** |  |  |
| Asymptomatic | 1 |  |
| Symptomatic | 1; (0.8-1.2) | 0.785 |
| **Occupation** |  |  |
| Doctor | 1 |  |
| Nurse | 1.0 (0.8-1.4) | 0.809 |
| Lab Technician | 1.3 (0.9-1.9) | 0.131 |
| Administration | 1.01 (0.8-1.5) | 0.766 |
| Others | 1.3(0.9-1.7) | 0.150 |

Association with seropositivity was further tested for correlation with gender, age, contact, morbidity, previous COVID symptoms, and occupation using both bivariate and multivariate analyses. However, only previous history of contact with confirmed or suspected COVID-19 case [COR 1.5 95% (1.3-1.9; p=0.0001) and AOR 1.4 (1.1-1.8; p=0.015)] and having symptoms compatible with COVID-19 in preceding 4 weeks [COR 1.3 (1.0-1.5] were found to be associated with seropositivity (Table 2).

## 4. Discussion

Interpretation of SARS-CoV-2 serologic test results, except pan Igs Wanti ELISA, has been reported to be very challenging in Africa due to pre-existing cross-reactive antibodies induced by other pathogens such as non-SARS-CoV-2 human coronaviruses and malaria parasites^14^. Given the rapid decline of anti-SARS-CoV-2 nucleocapsid antibodies as compared to the anti-RBD IgG antibody^13^, we developed and optimized an in-house ELISA that detects anti-SARS-CoV-2 IgG antibodies. Our assay, unlike other commercially available serologic assays, is affordable and has been validated with a large number of Ethiopian sera from both pre-COVID-19 and COVID-19 patients from the same regions. Its sensitivity on convalescent sera from COVID-19 patients confirmed by RT-PCR was found to be as sensitive as the Wantai pan Ig ELISA (100%), and superior to Realy Tech’s IgM/IgG LFA (90%). Also, our in-house assay displayed 97.7% specificity in randomly selected pre-COVID-19 Ethiopian origin sera, which is superior to Realy Tech (92.5 %).

Seroprevalence studies provide information about the extent of individuals who had exposure to to the virus and help to understand the future course of the pandemic and are key to providing target prevention and control measures in reducing transmission and severe outcomes^15^. In this study, the overall seroprevalence of SARS-CoV-2 spike RBD IgG antibodies among HWs was 39.6%, ranging from 24.5% in the Hiwot Fana Specialized Hospital, Harar to 48·0% in ALERT Hospital located in the capital city, Addis Ababa. This is not a surprise given Addis Ababa is the epicenter of SARS-CoV-2 transmission in Ethiopia, and SARS-CoV-2 has been introduced 4 months later in Harar. As a result of which, it is expected that a higher proportion of HWs in hospitals located in Addis Ababa, including ALERT are frequently exposed to COVID-19 cases than that HWs working in hospitals located in Harar, where fewer number cases and deaths had been reported.

According to our finding, at least 4 in 10 urban Ethiopian HWs had already been exposed to SARS-CoV-2 by February 2021 in Ethiopia. This result contrasts with a serosurvey in asymptomatic individuals from the general population conducted in March 2020 in Addis Ababa (8.8%)^16^ and from the household serosurveys in Jimma (2%) and Addis Ababa (5%) that were conducted during the first wave of the pandemic-i.e., four months after the first COVID-19 case in Ethiopia^17^. Although this stark seroprevalence difference between our study and these two previous studies might be explained by differences in the types of assays employed, lack of personal protective equipment (PPE) and/or respective cohorts, the most plausible explanation is that the sera for the present serosurveillance study had been collected after the first wave of the pandemic in Ethiopia, between March 2020 and February 2021.

While the high seroprevalence rates observed among the different geographically located hospitals are approaching those of high-incidence countries like Brazil^18^, they are in agreement with several other SARS-CoV-2 seroprevalence studies from sub-Saharan Africa that, like Ethiopia, have reported much lower rates of RT-PCR confirmed cases and deaths. For example, higher anti-SARS-CoV-2 antibody seroprevalence has been reported in South Sudan (30% to 60.6%) ^19^, Democratic Republic of Congo (8%-36)^20^ and Nigeria (25%-45) ^21^ depending on the population sampled and the serological test use. Taken together these studies indicate that SARS-CoV-2 has spread widely in sub-Saharan Africa ^22^. However, the majority (74.0%) of our study participants never had any symptoms compatible with COVID-19, suggesting the occurrence of significant burden of asymptomatic infections and its transmissions in the country, which is now, being reflected in the trend of increasing PCR positivity since January 2021. The higher proportion of younger HWs (mean age of 34 years), and the fewer participants with comorbidities (6.7%) may have contributed to the observed high burden of asymptomatic infection among the studied HWs. Malaria, BCG-vaccination, warmer environment, and high prevalence of pre-existing cross-reactivate against HCoVs may have also contributed^3^.

A report from Spain showed a higher (38.3%) seroprevalence of SARS-CoV-2 among HWs ^23^. This is comparable with the present report from Ethiopia, where there were a relatively fewer severe cases and deaths. Similarly, higher seroprevalence among frontline HWs has been reported in other sub-Saharan African countries such as in Malawi ^24^. These findings and ours highlight the importance of asymptomatic infections in the African countries. Interestingly, we found no seroprevalence differences between healthcare occupations including administrative staff. The lack of a dramatic difference between front line HWs and administrators may be a reflection of the frontline administrative staff are also at high risk and are poorly protected, or may suggest the level of virus transmission in the general population at large as previously observed in UK^9^. Nevertheless, further well-designed investigations are required to implement occupation-specific public health strategies in healthcare facilities.

In the present study, a history of previous close contact with a suspected or confirmed COVID-19 case was found to be strongly associated with seropositivity; however, this finding contradicts the observed similar seropositivity between front line HWs and administrators. Similar odds of seropositivity between males and females were also found although several studies elsewhere reported higher odds of seropositivity in males ^25^. A similar contradictory finding was reported in the Spanish general population^23^.

Our study has several strengths. These include its use of an in-house developed assay which we optimized to significantly minimize false positive responses by validating it with both pre-pandemic and pandemic samples of Ethiopian origin. Most importantly, the study involved a relatively large sample size from five hospitals located in different geographical locations, providing much needed information about the COVID-19 pandemic in sub-Saharan Africa.

Despite these strengths, our study has several limitations. First, all hospital staff were invited to take part in the study, and hence selection bias might have affected our results. Second, recall bias might have affected the responses to the history of symptoms compatible with COVID-19, and close contact with a confirmed COVID-19 case, and thereby contributed to the absence of a strong correlation between seropositivity and these covariates, albeit having close contact with COVID-19 case. Third, our findings are slightly affected by the accuracy of our assay, with a sensitivity of 100% in convalescent samples from RT-PCR conformed COVID-19 cases and specificity of 97.7% in pre-COVID-19 samples.

However, even this slight overestimation of the apparent seroprevalence associated with the assay specificity is likely to be matched by the proportion of study participants who might be infected and yet not produce humoral immune responses at the time of blood sample collection.

In conclusion, we developed an in-house IgG ELISA that meets the WHO requirements to be utilized for SARS-CoV-2 serosurveillance studies. This seroprevalence study revealed a remarkably high seroprevalence (40-48%) of SARS-CoV-2 among HWs in the five public hospitals; with slight differences amongst hospitals, except Hiwot Fana Specialized Hospital in which relatively lowest (24.5%) seroprevalence was found. We found no seroprevalence rate differences between front line HWs and administrative staff, indicating the observed high seroprevalence of SARS-CoV-2 might also be a reflection of the community transmission. Taken together these findings suggest extensive cryptic circulation (asymptomatic transmission) of SARS-CoV-2 in Ethiopia. Whether the detected anti-SARS-CoV-2 antibodies can persist adequately and confer protection from subsequent infections to those HWs who had or had not received COVID-19 vaccine will require further immunological investigation.

## Data Availability

All the data are included in the manuscript

## Acknowledgements

We thank study participants and AHRI’s SARS-CoV-2 diagnostic and research team members. We also thank Mr. Mekonnen Ashagarie, Director of American Health and Home Care, MA, USA for the non-technical support.

## Contributors

TG, AM, AMi MA, AA and FG conceived the study. TG, BS, AM, and AA wrote the first draft of the protocol, and revised by all authors. TG, AM, MA, AA and FGT developed the serologic assay, TAB purified the antigen. TG, BS, AM, AMi and AA coordinated the sample collection. BG, AS, YM, YM, ZT, DK, AG, DA and ET collected the blood samples and data. BT, MO, GJ, AA, AH and DT conducted the sample testing, TH cleaned the data, TH and TG analysed the data, TG accessed and verified the data underlying the study and take responsiblity for the data. TG and AA drafted the manuscript. All authors edited the final manuscript. All authors contributed to study design, revising the protocol and the manuscript for important intellectual content. TG, AA, FGT were responsible for the decision to submit for publication, and approved the final submitted version of the manuscript.

---

**Table S1.** The Specificity of RBD IgG ELISA among pre-COVID-19 pandemic sera (n=365)

| Negative Samples | Total number tested | Negative | Specificity (%) | 95% CI |
| --- | --- | --- | --- | --- |
| Pre-COVID-19 pandemic sera | 364 | 350 | 97.7 | 95.6-99.0 |
| NIBS UK adults pooled plasma | 1 | 1 |  |  |
| Total | 365 | 351 | 97.7 | 95.6-99.0 |

**Table S2.** Sensitivity of RBD IgG ELISA among cohort COVID-19 patients (n=405) confirmed by RT-PCR.

| Sample collection | Total N Tested | Positive | Sensitivity (%) | 95% CI |
| --- | --- | --- | --- | --- |
| 1-7dps | 336 | 226 | 67.3 | 62.0-72.3 |
| 8-14dps | 52 | 39 | 75.0 | 61.0-86.0 |
| 15-21dps | 13 | 13 | 100.0 | 75.29-100.0 |
| NIBSC UK COVID-19 convalescent plasma | 4 | 4 | 100.0 | - |
| Total Convalescent samples s | 17 | 17 | 100.0 | 84.2-100 |

**Table S3.**
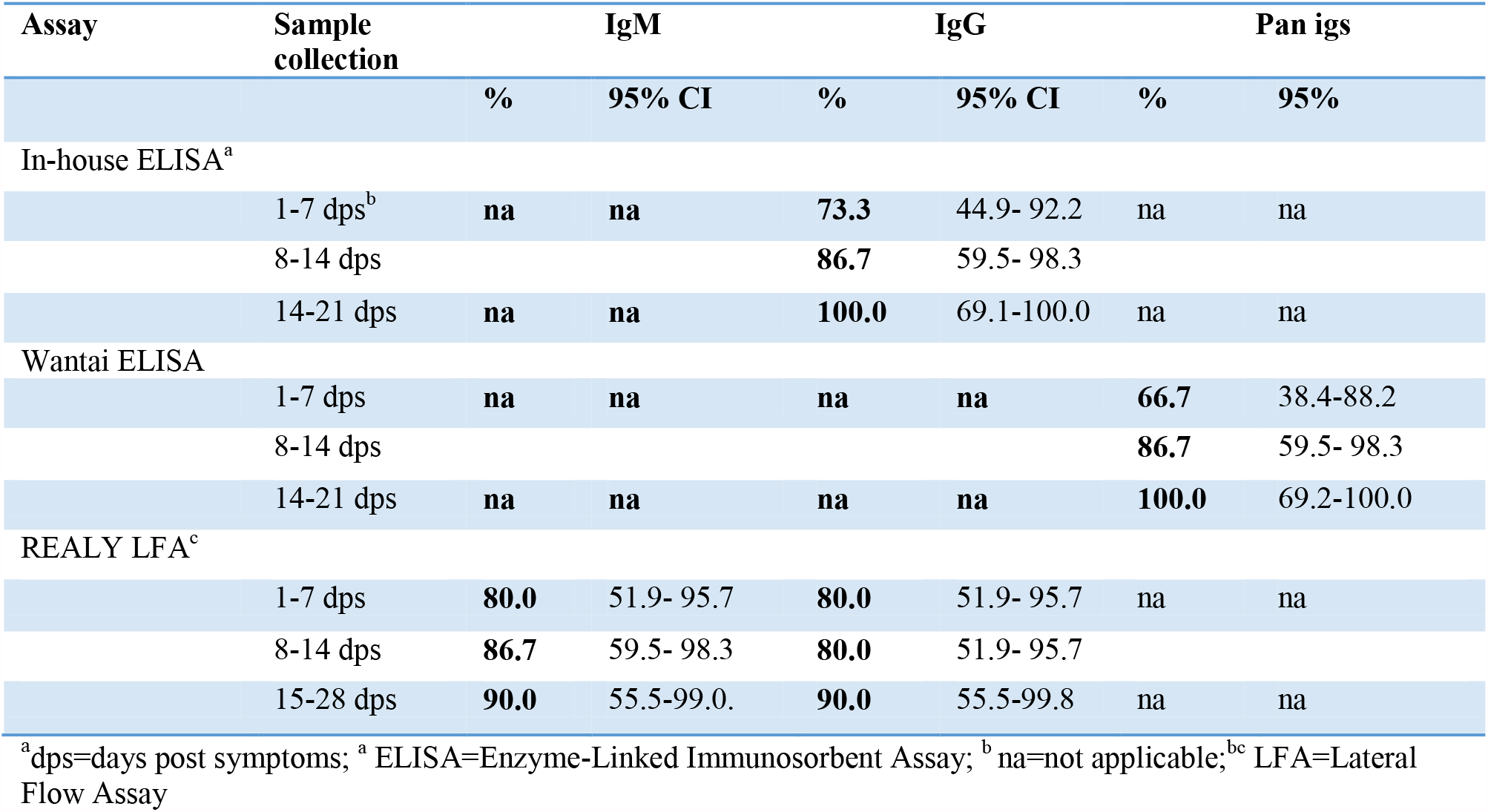
Percentage of positive specimens (n=40) from patients who tested positive for SARS-CoV-2 by DAAn RT–PCR.

| Assay | Sample collection | IgM |  | IgG |  | Pan igs |  |
| --- | --- | --- | --- | --- | --- | --- | --- |
|  |  | % | 95% CI | % | 95% CI | % | 95% |
| In-house ELISA<sup>a</sup> |  |  |  |  |  |  |  |
|  | 1-7 dps<sup>b</sup> | na | na | 73.3 | 44.9- 92.2 | na | na |
|  | 8-14 dps |  |  | 86.7 | 59.5- 98.3 |  |  |
|  | 14-21 dps | na | na | 100.0 | 69.1-100.0 | na | na |
| Wantai ELISA |  |  |  |  |  |  |  |
|  | 1-7 dps | na | na | na | na | 66.7 | 38.4-88.2 |
|  | 8-14 dps |  |  |  |  | 86.7 | 59.5- 98.3 |
|  | 14-21 dps | na | na | na | na | 100.0 | 69.2-100.0 |
| REALY LFA<sup>c</sup> |  |  |  |  |  |  |  |
|  | 1-7 dps | 80.0 | 51.9- 95.7 | 80.0 | 51.9- 95.7 | na | na |
|  | 8-14 dps | 86.7 | 59.5- 98.3 | 80.0 | 51.9- 95.7 |  |  |
|  | 15-28 dps | 90.0 | 55.5-99.0. | 90.0 | 55.5-99.8 | na | na |

<sup>a</sup>dps=days post symptoms; <sup>a</sup> ELISA=Enzyme-Linked Immunosorbent Assay; <sup>b</sup> na=not applicable; <sup>bc</sup> LFA=Lateral Flow Assay

**Table S4.**
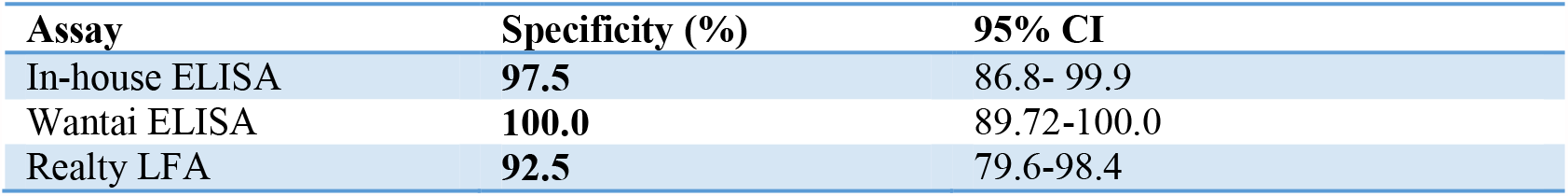
Specificity of RBD IgG ELISA in pre-covid plasma/serum specimens (n=40) collected before COVID-19 pandemic.

## Supplementary Method

Microtiter plates were coated with purified recombinant proteins of receptor binding domain of the spike protein of SARS-CoV-2 (100 μl/well) diluted in phosphate buffered saline, PBS (pH 7.4) at concentration 1 μg/mL and incubated overnight at 4 °C. Next day, excess unbound antigen was removed and thereafter microtiter plates were blocked with 300 μl/well of 4% skimmed milk with PBS plus 0.1% Teween-20 (w/v) for 2 hours at room temperature (RT). Following blocking step, microtiter plates washed 3X with PBS plus 0.05% Tween-20 (PBST) and thereafter 100 μl/ml of serum sample diluted at 1:200 in blocking buffer was added and incubated at RT for 60 min. Following incubation and 5X washes with PBST, 100 μl/well of horseradish peroxidase-conjugated anti-human immunoglublunin G (IgG) (Invitrogen, USA) diluted at 1:5000 in blocking buffer was added and incubated for 1 h at RT. After 5X washes, the reaction was visualized by adding 75 μl/well 3,3_,5,5_-Tetramethylbenzidine (TMB) liquid substrate (BioRad, USA) and incubating at RT in the dark for 10-15 min. The reaction was then stopped with 75 μl/well TMB stop solution. The optical density (OD) was measured at 450 nm filter on ELISA LT-45000 microplate reader. Each sample was tested in duplicate.

## References

1 Coronavirus Disease 2019 (COVID-19) – Africa CDC. https://africacdc.org/covid-19/ (accessed March 18, 2021).

2 Gaye B, Khoury S, Cene CW, et al. Socio-demographic and epidemiological consideration of Africa’s COVID-19 response: what is the possible pandemic course? Nat Med 2020; 26: 996–9.

3 Tso FY, Lidenge SJ, Peña PB, et al. High prevalence of pre-existing serological cross-reactivity against severe acute respiratory syndrome coronavirus-2 (SARS-CoV-2) in sub-Saharan Africa. Int J Infect Dis 2021; 102: 577–83.

4 Yitbarek K, Abraham G, Girma T, Tilahun T, Woldie M. The effect of Bacillus Calmette-Guérin (BCG) vaccination in preventing severe infectious respiratory diseases other than TB: Implications for the COVID-19 pandemic. Vaccine 2020; 38: 6374–80.

5 Kalungi A, Kinyanda E, Akena DH, Kaleebu P, Bisangwa IM. Less Severe Cases of COVID-19 in Sub-Saharan Africa: Could Co-infection or a Recent History of Plasmodium falciparum Infection Be Protective? Front Immunol 2021; 12: 565625.

6 Verity R, Okell LC, Dorigatti I, et al. Estimates of the severity of coronavirus disease 2019: a model-based analysis. Lancet Infect Dis 2020; 20: 669–77.

7 WHO-2019-nCoV-Seroepidemiology-2020.2-eng.pdf. https://apps.who.int/iris/bitstream/handle/10665/332188/WHO-2019-nCoV-Seroepidemiology-2020.2-eng.pdf?sequence=1&isAllowed=y (accessed March 18, 2021).

8 Garcia-Basteiro AL, Moncunill G, Tortajada M, et al. Seroprevalence of antibodies against SARS-CoV-2 among health care workers in a large Spanish reference hospital. Nat Commun 2020; 11: 3500.

9 Treibel TA, Manisty C, Burton M, et al. COVID-19: PCR screening of asymptomatic health-care workers at London hospital. Lancet Lond Engl 2020; 395: 1608–10.

10 ’Solidarity 2’ global serologic study for COVID-19. https://www.who.int/emergencies/diseases/novel-coronavirus-2019/global-research-on-novel-coronavirus-2019-ncov/solidarity-2-global-serologic-study-for-covid-19 (accessed March 18, 2021).

11 Isho B, Abe KT, Zuo M, et al. Persistence of serum and saliva antibody responses to SARS-CoV-2 spike antigens in COVID-19 patients. Sci Immunol 2020; 5. DOI:10.1126/sciimmunol.abe5511.

12 Bates TA, Weinstein JB, Farley S, Leier HC, Messer WB, Tafesse FG. Cross-reactivity of SARS-CoV structural protein antibodies against SARS-CoV-2. Cell Rep 2021; 34: 108737.

13 openFDA. https://open.fda.gov/apis/device/covid19serology/ (accessed April 15, 2021).

14 Emmerich P, Murawski C, Ehmen C, et al. Limited specificity of commercially available SARS-CoV-2 IgG ELISAs in serum samples of African origin. Trop Med Int Health TM IH 2021; published online March 5. DOI:10.1111/tmi.13569.

15 Clapham H, Hay J, Routledge I, et al. Seroepidemiologic Study Designs for Determining SARS-COV-2 Transmission and Immunity. Emerg Infect Dis 2020; 26: 1978–86.

16 Alemu BN, Addissie A, Mamo G, et al. Sero-prevalence of anti-SARS-CoV-2 Antibodies in Addis Ababa, Ethiopia. bioRxiv 2020; : 2020.10.13.337287.

17 Abdella S, Riou S, Tessema M, et al. Prevalence of SARS-CoV-2 in urban and rural Ethiopia: Randomized household serosurveys reveal level of spread during the first wave of the pandemic. EClinicalMedicine 2021; 35. DOI:10.1016/j.eclinm.2021.100880.

18 Buss LF, Prete CA, Abrahim CM, et al. COVID-19 herd immunity in the Brazilian Amazon. medRxiv2020; : 2020.09.16.20194787.

19 Wiens KE, Mawien PN, Rumunu J, et al. Seroprevalence of anti-SARS-CoV-2 IgG antibodies in Juba, South Sudan: a population-based study. MedRxiv Prepr Serv Health Sci 2021; published online March 12. DOI:10.1101/2021.03.08.21253009.

20 Mukwege D, Byabene AK, Akonkwa EM, et al. High SARS-CoV-2 Seroprevalence in Healthcare Workers in Bukavu, Eastern Democratic Republic of Congo. Am J Trop Med Hyg 2021; 104: 1526–30.

21 SARS-CoV-2 Seropositivity in Asymptomatic Frontline Health Workers in Ibadan, Nigeria in: The American Journal of Tropical Medicine and Hygiene Volume 104 Issue 1 (2020). https://www.ajtmh.org/view/journals/tpmd/104/1/article-p91.xml (accessed April 13, 2021).

22 Usuf E, Roca A. Seroprevalence surveys in sub-Saharan Africa: what do they tell us? Lancet Glob Health 2021; 0. DOI:10.1016/S2214-109X(21)00092-9.

23 Prevalence of SARS-CoV-2 in Spain (ENE-COVID): a nationwide, population-based seroepidemiological study - The Lancet. https://www.thelancet.com/journals/lancet/article/PIIS0140-6736(20)31483-5/fulltext (accessed April 13, 2021).

24 High SARS-CoV-2 seroprevalence in health care workers but relatively low numbers of deaths in urban Malawi |medRxiv. https://www.medrxiv.org/content/10.1101/2020.07.30.20164970v3 (accessed April 13, 2021).

25 Galanis P, Vraka I, Fragkou D, Bilali A, Kaitelidou D. Seroprevalence of SARS-CoV-2 antibodies and associated factors in healthcare workers: a systematic review and meta-analysis. J Hosp Infect 2021; 108: 120–34.

